# Evolving Phenotypes of non-hospitalized Patients that Indicate Long Covid

**DOI:** 10.1101/2021.04.25.21255923

**Authors:** Hossein Estiri, Zachary H Strasser, Gabriel A Brat, Yevgeniy R Semenov, The Consortium for Characterization of COVID-19 by EHR (4CE), Chirag J Patel, Shawn N Murphy

**Affiliations:** Laboratory of Computer Science, Massachusetts General Hospital, Boston, MA, 02144, USA; Department of Medicine, Massachusetts General Hospital, Boston, MA, 02144, USA; Department of Biomedical Informatics, Harvard Medical School, Boston, MA, USA; Department of Dermatology, Massachusetts General Hospital, Boston, MA, 02114, USA; Department of Neurology, Massachusetts General Hospital, Boston, MA, 02114, USA; Research Information Science and Computing, Mass General Brigham, Boston MA, USA

## Abstract

For some SARS-CoV-2 survivors, recovery from the acute phase of the infection has been grueling with lingering effects. Many of the symptoms characterized as the post-acute sequelae of COVID-19 (PASC) could have multiple causes or are similarly seen in non-COVID patients. Accurate identification of phenotypes will be important to guide future research and help the healthcare system focus its efforts and resources on adequately controlled age- and gender-specific sequelae of a COVID-19 infection. In this retrospective electronic health records (EHR) cohort study, we applied a computational framework for knowledge discovery from clinical data, MLHO, to identify phenotypes that positively associate with a past positive reverse transcription-polymerase chain reaction (RT-PCR) test for COVID-19. We evaluated the post-test phenotypes in two temporal windows at 3-6 and 6-9 months after the test and by age and gender. Data from longitudinal diagnosis records stored in EHRs from Mass General Brigham in the Boston metropolitan area was used for the analyses. Statistical analyses were performed on data from March 2020 to June 2021. Study participants included over 96 thousand patients who had tested positive or negative for COVID-19 and were not hospitalized. We identified 33 phenotypes among different age/gender cohorts or time windows that were positively associated with past SARS-CoV-2 infection. All identified phenotypes were newly recorded in patients’ medical records two months or longer after a COVID-19 RT-PCR test in non-hospitalized patients regardless of the test result. Among these phenotypes, a new diagnosis record for anosmia and dysgeusia (OR: 2.60, 95% CI [1.94 - 3.46]), alopecia (OR: 3.09, 95% CI [2.53 - 3.76]), chest pain (OR: 1.27, 95% CI [1.09 - 1.48]), chronic fatigue syndrome (OR 2.60, 95% CI [1.22-2.10]), shortness of breath (OR 1.41, 95% CI [1.22 - 1.64]), pneumonia (OR 1.66, 95% CI [1.28 - 2.16]), and type 2 diabetes mellitus (OR 1.41, 95% CI [1.22 - 1.64]) are some of the most significant indicators of a past COVID-19 infection. Additionally, more new phenotypes were found with increased confidence among the cohorts who were younger than 65. Our approach avoids a flood of false positive discoveries while offering a more robust probabilistic approach compared to the standard linear phenome-wide association study (PheWAS). The findings of this study confirm many of the post-COVID symptoms and suggest that a variety of new diagnoses, including new diabetes mellitus and neurological disorder diagnoses, are more common among those with a history of COVID-19 than those without the infection. Additionally, more than 63 percent of PASC phenotypes were observed in patients under 65 years of age, pointing out the importance of vaccination to minimize the risk of debilitating post-acute sequelae of COVID-19 among younger adults.

## Background

The onslaught of the COVID-19 pandemic in the United States and around the world was relentless. For hundreds of thousands (if not millions), recovery from the acute phase of the SARS-CoV-2 infection, the coronavirus that causes COVID-19, will be grueling with a debilitating second act. A collection of persistent physical (e.g., fatigue, dyspnea, chest pain, cough), psychological (e.g., anxiety, depression, post-traumatic stress disorder), and neurocognitive symptoms (e.g., impaired memory and concentration) can appear and last for weeks or months in patients after acute COVID-19.[1–8] Many of the symptoms characterized as the post-acute sequelae of COVID-19 (PASC) could have multiple causes.

So far, a number of studies have been published on PASC,[1–7, 9, 10] but most have small samples, case-series, or rely on self-reports. Carfi et al assessed 179 hospitalized COVID patients in Italy at an average of 60 days after the onset of symptoms using a standard questionnaire.[11] Only 12.6% were completely free of all COVID-19 symptoms and 55% had 3 or more symptoms. The most common symptoms were fatigue, dyspnea, joint pain, and chest pain. Chopra et al performed an observational study of 488 patients who were hospitalized 60 days after their discharge with a phone survey.[12] The most common persistent symptoms were cough, dyspnea, persistent loss of taste or smell, and worsening difficulty completing activities of daily living. Huang et al performed one of the larger cohort studies where they analyzed 1,733 COVID patients discharged from a hospital in China with a questionnaire at 6 months.[13] They identified fatigue, muscle weakness, sleep difficulties, anxiety, and depression as the most common symptoms 6 months after the initial diagnosis.

These studies are all case series, focusing only on patients with COVID-19. Additionally, prior PASC studies often focus on patients with severe COVID-19 symptoms after hospitalization. It is unclear whether the identified persistent symptoms hold true among Covid patients not hospitalized. Furthermore, many of the published studies are based on small cohorts (several hundred COVID-19 patients were analyzed) and relied on self-reported outcomes which can embody potential biases due to, for example, exaggeration of symptoms.[14]

There have also been a number of less commonly reported symptoms including ocular inflammation[15] cardiac involvement, [16, 17] autonomic instability,[18] recurrent pseudomonas infections,[19] persistent mucous secretion,[20] micro-structural changes to the brain[21] and Guillain-Barre syndrome.[22] A large cohort analyzing the ICD-10 (the 10th revisions of the International Statistical Classification of Diseases and Related Health Problems) diagnoses in the electronic health record between patients with and without a history of COVID could help clarify the actual association with the disease.

We present results from a retrospective cohort study of over 97,000 patients with an RT-PCR test for COVID-19 in a Mass General Brigham (MGB) facility. We detected de novo phenotypes that appeared for the first time in EHRs at two temporal windows of 3-6 and 6-9 months after a COVID-19 test for both COVID-positive and -negative patients. Leveraging MLHO, a computational framework developed for knowledge discovery from electronic health records (EHRs)[23–25] with a validated utility for studying and modeling post-COVID outcomes[26, 27] augmented with clinical expertise, we identified 33 phenotypes in different age/gender groups or time windows positively associated with a recent/past SARS-CoV-2 infection. All identified phenotypes were newly recorded in patients’ medical records two months or longer after a COVID-19 RT-PCR test in non-hospitalized patients regardless of the test result.

## Methods

We utilized longitudinal EHR diagnosis records from all patients who tested for SARS-CoV-2 infection -- reverse transcription-polymerase chain reaction (RT-PCR) -- between March 2020 and June 2021 in a Mass General Brigham (MGB) facility. We limited the patient cohort to those who were alive and not hospitalized. To increase the confidence that a patient in our cohort would likely seek care within MGB in the post-COVID era, we further narrowed the study population to patients who had two diagnosis records, 6 months apart, in our electronic data repositories since 2010. We also excluded patients who had a diagnosis code referring to past COVID-19 but having a negative RT-PCR test in the MGB records due to our inability to approximate the infection date. The use of clinical data in this study was approved by the MGB Institutional Review Board with a waiver of informed consent.

### Phenotype coding

To construct the feature space, we utilized EHR diagnoses recorded in ICD-9 and ICD-10 codes (the 9th and 10th revisions of the International Statistical Classification of Diseases and Related Health Problems). To represent phenotypes for the analyses, we mapped the ICD-9/10 diagnosis codes to a unique phenotype code (PheCode) from the Phenome-wide association studies (PheWAS)[28, 29] groups of phenotypes. We assigned a temporal buffer of two months after the RT-PCR test as a proxy for the acute phase in COVID-19 patients and used the first observation of phenotypes that were recorded for the first time after the acute phase (Figure 1). Using this temporal segmentation, we further limited the data, by only using the first observation of the records (to minimize the problem list repetitions) and only considered the diagnosis records that for the first time appeared in a patient’s medical records two months or longer after the RT-PCR test -- see eMethods for more details. As such, the feature space contained all PheCodes that were recorded for the first time in a patient’s longitudinal EHR data two months or later after the COVID-19 RT-PCR test, regardless of the test result.

**Figure 1.**
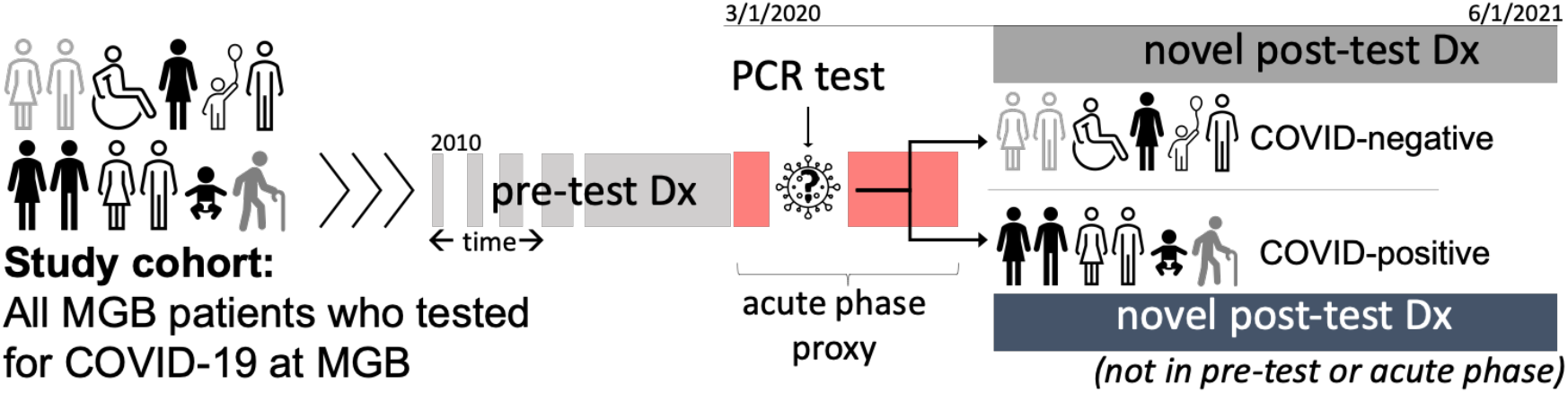
Study cohort, temporal segmentation, and diagnoses recording.

### MLHO framework

To robustly identify the phenotypes that are positively associated with a recent positive test for COVID-19, we applied a multivariate temporal approach to classifying past RT-PCR test results from the post-test clinical data. The classification algorithm here is not intended for the purpose of classification. Rather, we performed “ postdiction,” which is the “ assertion or deduction about something in the past,” [30] aiming to identify features (i.e., phenotype) that carry information to make such an assertion about the past event. To do so, we leveraged the MLHO framework,[26] which includes a suite of computational algorithms[23, 26] specifically designed for modeling and phenotyping clinical data. We followed a similar analytic process used by Estiri et al. (2021)[31] that was used to identify risk factors for COVID-19 mortality from EHR data. From the MLHO framework, the computational process involved applying the Minimize Sparsity, Maximize Relevance (MSMR) algorithm,[23, 32, 33] clinical expertise, and multivariate boosting logistic regression, to compute a composite confidence score for identifying the phenotypes that are positively associated with a past RT-PCR test (see eMethods for more details).

All analyses were conducted in R statistical language.

### Cohort stratification

To increase specificity, we stratified the analyses by age and gender in a nested structure. This resulted in the following strata: 1) all patients, 2) 65 and older, 3) under 65, 4) 65 and older female, 5) 65 and older male, 6) under 65 female, and 7) under 65 male. In addition to stratifying the cohort, we controlled for the age and gender (in gender-agnostic models) of the patient. For the phenotypes identified by MLHO in each stratified model, we trained standard generalized logistic regression models controlling for age and gender and extracted multivariate Odds Ratios (ORs) along with p-value (Wald’s test) and 95% confidence intervals using profiled log-likelihood.

### Clinical validation via chart reviews

Due to the known reliability issues of EHR diagnosis records,[33, 34] we validated the phenotypes identified by MLHO through chart reviews. A clinical expert reviewed the clinical notes and longitudinal records for a random sample of five patients for each phenotype identified by MLHO with an 80-plus confidence score. The chart review required reviewing the clinical notes at the time of the diagnostic code to determine whether the phenotype was actually present at the encounter and whether this was a new symptom or diagnosis since the time of the COVID encounter. If at least three of the randomly sampled five charts verified the phenotype’s presence and its recent appearance or diagnosis, then the phenotype was included in the final analysis. Two phenotypes were

## Results

From over 397,000 patients who tested for COVID-19 in an MGB facility with a nasal swab, 210,949 met our inclusion/exclusion criteria, including 52,491 patients with positive test results. After applying the approach for keeping records, 96,025 patients remained in our final study cohort, 22,475 (23.41%) of whom were positive for the SARS-CoV-2 virus (Table 1S in eAppendix and Figure 2S in eMethods). After the sparsity screening (i.e., removing low prevalence [<0.22%] phenotypes from sub-cohorts), 354 and 334 phenotypes were evaluated in the full cohorts during the 3-6 and 6-9 month temporal windows.

**Figure 2.**
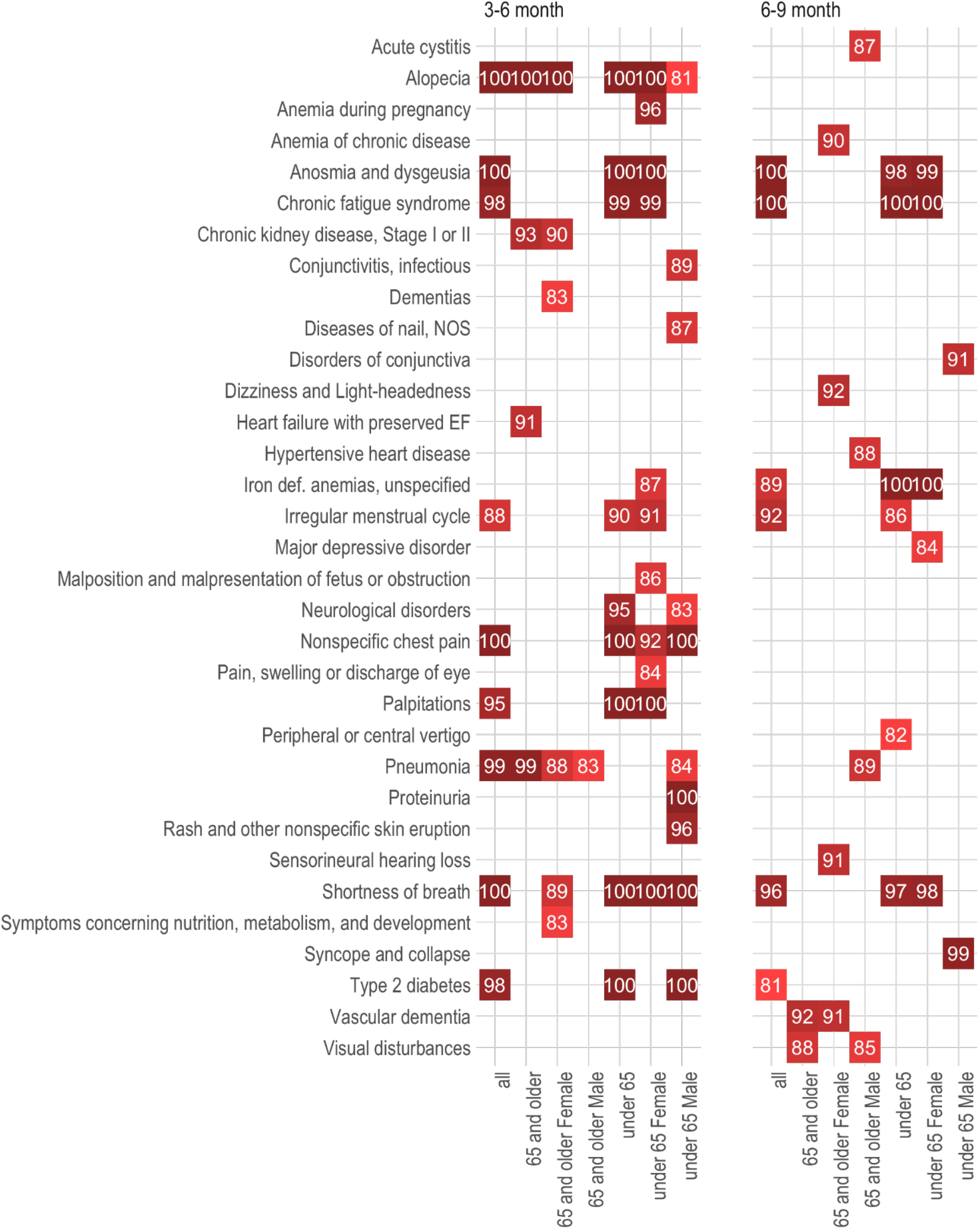
Phenotypes that are positively associated with a past COVID-19 positive RT-PCR test. * MLHO confidence scores are plotted in white font. 100% means identified in all 100 MLHO iterations. Phenotypes included have been associated with a positive past COVID-19 test with a confidence score higher than 80 percent in at least a sub-cohort.

Overall, MLHO identified 41 phenotypes in different age/gender groups and/or time windows as positively associated with a past positive COVID-19 test, with a MLHO confidence score higher than 80. All identified phenotypes were newly recorded in patients’ medical records two months or longer after a COVID-19 RT-PCR test in non-hospitalized patients regardless of the test result. We performed chart reviews on 215 randomly sampled patients to validate MLHO’s findings. For nearly all of the phenotypes, the details and descriptions provided in the clinical notes matched with the assigned phenotype for that chart (eAppendix Table S3). For 33 of the phenotypes (Figures 2 and 3), the majority of the random samples of notes reviewed were suggestive that the phenotype was new since the time of COVID. Accordingly, we removed 8 phenotypes due to the likelihood they were present pre-COVID based on the notes, despite the use of a new ICD-9/10 record since the COVID-19 diagnosis. For the 33 phenotypes, multivariate Odds Ratios (ORs), 95 percent confidence intervals, and MLHO’s Confidence Scores (CSs) are provided below -- also available in table 2S in eAppendix.

**Figure 3.**
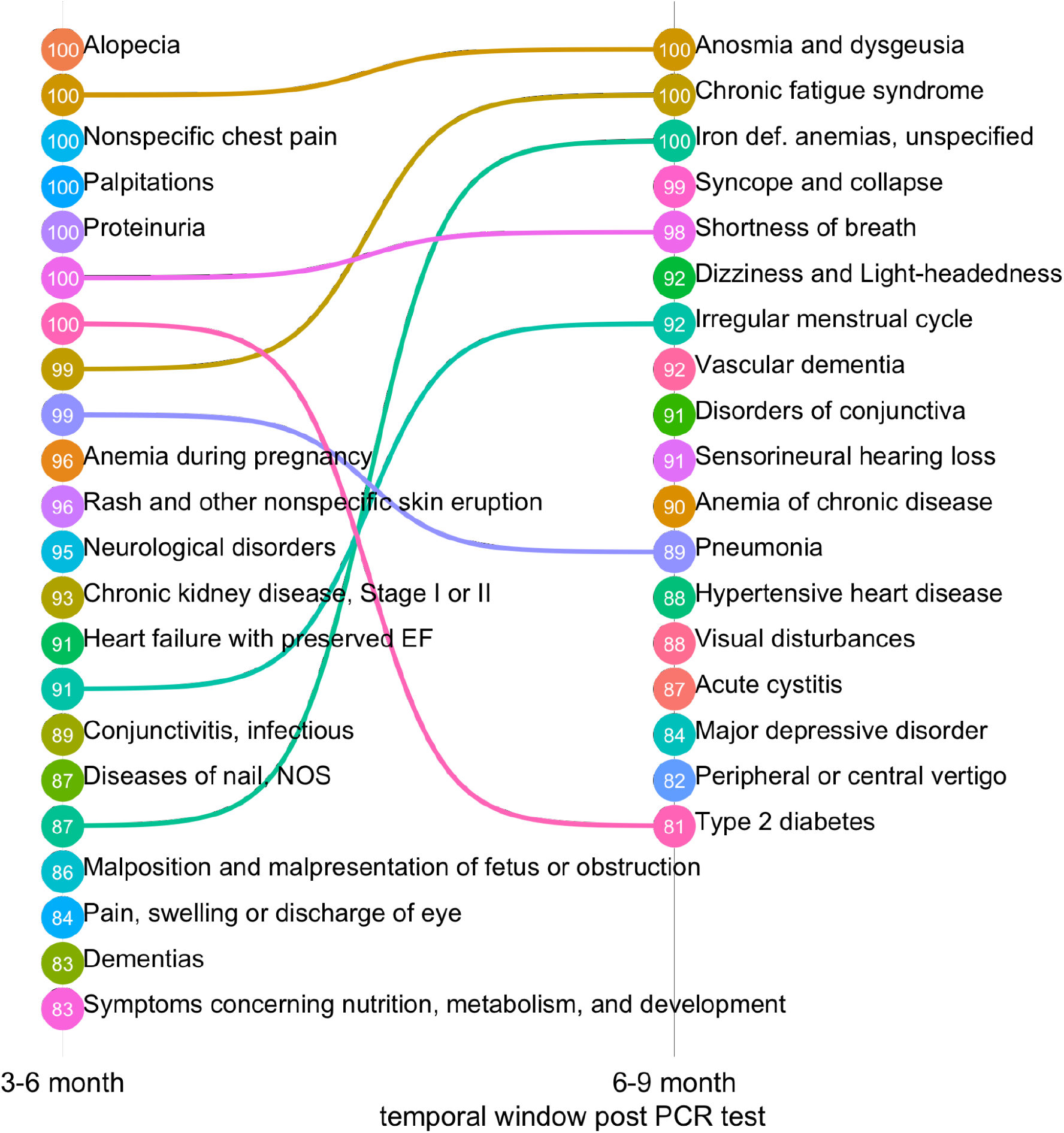
Presentation of the PASC phenotypes across temporal windows. * phenotypes are ranked by MLHO confidence score.

Results demonstrated extremely high confidence (>97%) in eleven phenotypes, which in the overall cohort and/or one or more sub-cohorts indicate a positive past COVID-19 infection. Seven were very high among the entire population in the 3-6 month window. Alopecia was identified in all iterations of MLHO between months three and six, in the overall cohort (OR: 3.09, 95% CI [2.53 - 3.76], CS: 100). It was also specifically seen in those younger and older than 65 cohorts, and specifically in women both under and over 65. Similarly, a new diagnosis record of nonspecific chest pain was indicative of past COVID-19 infection in the 3-6 month temporal window (OR: 1.27, 95% CI [1.09 - 1.48], CS: 100), and particularly among people under 65 (OR: 1.30, 95% CI [1.08 - 1.55], CS: 100). Anosmia and dysgeusia were identified in 100 percent of the MLHO iterations, in the 3-6 month window (OR: 2.60, 95% CI [1.94 - 3.46], CS: 100) and continued to be important in the 6-9 month window (OR: 2.10, 95% CI [1.40 - 3.11], CS:100). The phenotype was indicative of past positive COVID-19 in those under 65, and women under 65.

Among other identified phenotypes with 97 and higher confidence scores, was chronic fatigue syndrome seen in both the 3-6 month window (OR 2.60, 95% CI [1.22-2.10], CS: 98) and the 6-9 month window (OR 2.03, 95% CI [1.31-3.11]), and appearing more prominent in the patients less than 65 and women less than 65. Pneumonia, in the 3-6 month window, had a high confidence score among the overall population (OR 1.66, 95% CI [1.28 - 2.16], CS: 99) and those older than 65 (OR 1.92, 95% CI [1.03-3.46], CS: 99). Shortness of breath had high confidence scores in both the 3-6 month window (OR 1.41, 95% CI [1.22 - 1.64], CS 100) and the 6-9 month window (OR 1.45, 95% CI [1.09 - 1.93], CS 96). It also was identified as having a high confidence score among those under 65. Finally, palpitations (OR 1.41, 95% CI [1.22 - 1.64]) in the 3-6 month range and type 2 diabetes mellitus in the 3-6 (OR 1.41, 95% CI [1.22 - 1.64]) also had high confidence scores.

Several phenotypes had very high scores but only within certain time frames and in certain sub-cohorts. For example, iron-deficiency anemia in the 6-9 month range for those under 65 (OR 2.02, 95% CI [1.37 - 2.95], CS: 100) and women under 65 (OR 2.10, 95% CI [1.40 - 3.15], CS: 100). Men under 65 were identified with proteinuria (OR 3.19, 95% CI [1.72 - 5.96], CS: 100) in the 3-6 month range and syncope and collapse (OR 4.80, 95% CI [1.56 - 13.39], CS: 99) in the 6-9 month range.

Among other COVID-19 related phenotypes identified as indicators of past COVID-19 infection with a 90-to-96 confidence score were a number of subgroups. In the 3-6 month window, this includes anemia during pregnancy in women under 65, chronic kidney disease in the cohort older than 65 and women over 65, heart failure with preserved ejection fraction in the cohort older than 65, irregular menstrual cycle in women under 65, neurological disorders in those under 65, and rash and other nonspecific skin eruptions in men under 65. In the 6-9 month range phenotypes, with a confidence score in the 90-to-96 window, including anemia of chronic disease in women 65 and older, disorders of the conjunctiva in men under 65, dizziness and lightheadedness in women older than 65, irregular menstrual cycle in the total cohort, sensorineural hearing loss in women greater than 65, and vascular dementias for those older than 65 and women older than 65.

## Discussion

We identified 33 phenotypes that were indicative of long Covid among non-hospitalized COVID- 19 patients. Phenotypes such as alopecia, anosmia, fatigue, shortness of breath, and chest pain, have been well documented as common signs and symptoms of PASC.[7, 35, 36] This study shows that these phenotypes are some of the earliest associations with the syndrome seen in the 3-6 month window after the initial infection and some of the most important features for indicating previous COVID-19 infection. All five of these phenotypes (alopecia, anosmia and dysgeusia, shortness of breath, chronic fatigue syndrome, and nonspecific chest pain) were documented with high confidence in the 3-6 month window. And while alopecia and non-specific chest pain were not found with high confidence in the 6-9 month window, anosmia and chronic fatigue syndrome continued to be important phenotypes seen in both time periods. Additionally, several phenotypes were identified with similarly high confidence including type II diabetes, pneumonia, proteinuria, and syncope and collapse.

Interestingly, those aged less than 65 had more new phenotypes identified with greater confidence, than the cohorts who were older than 65. Over 63 percent of the identified long Covid phenotypes were observed in past COVID-19 patients who were under 65 years old. These findings have important implications for younger patients. Despite having not been hospitalized during the acute phase, the symptoms of long Covid are found with high confidence in this younger cohort population. This gives another reason for young patients to opt for having the vaccination since the long-term effects of the disease are clearly not limited to older patients. While the precise biological causes of the sequelae are still unknown and under investigation, the enrichment of these diagnoses among younger cohorts may indicate that the robustness of the immune response in these patients is driving some of the post-COVID sequelae. However, these results should be understood and qualified in the context that, on average, younger patients who are often healthier than 65 and older have fewer interactions with healthcare systems (and thus fewer diagnosis records), which may lead to greater ease in detecting a signal in this younger cohort compared to an older cohort.

While the chart review’s primary purpose was to determine if the clinical notes were in agreement with the ICD-9/10 labels, the reviewer also noted that physicians consistently attributed two of the phenotypes (alopecia, and anosmia and dysgeusia) to a previous history of COVID-19. Whereas, the physicians’ notes did not specifically identify a connection between the phenotype and the previous infection for most of the other phenotypes, even those with high confidence like type 2 diabetes or non-specific chest pain. Our model indicates that even if these phenotypes are not explicitly identified or recognized by the clinician and patient at the individual level, many of these unrecognized phenotypes still have a high confidence score. While an ICD code on its own does not specify the time of onset, the chart review helped to confirm that the presented phenotypes were likely new since COVID-19. The majority of charts reviewed for each phenotype suggest that the symptoms or the diagnosis occurred after COVID-19. Our model identifies relationships between COVID and a phenotype, where a healthcare provider and patient may otherwise miss that relationship.

Several neurological phenotypes (vascular dementia, dementia, and neurological disorders) were frequently diagnosed after COVID and appear to have an increased association with the infection. The earliest reports of acute COVID, such as Mao’s retrospective analysis of 214 hospitalized patients in China, described neurological manifestations, including cerebrovascular complications, in nearly half of those with severe disease.[37] Since the acute phase, the sequelae for the description of “ brain fog” after the diagnosis of COVID have been repeatedly described.[38, 39] Al-Aly specifically documents increased memory problems and strokes.[40] Our model suggests that in some cases these symptoms are so severe they are even leading to an initial formal diagnosis of dementia at higher rates among those with a history of COVID. While many of these patients may have already shown some signs of memory loss, the formal diagnosis of dementia didn’t come until after COVID-19 suggesting that the viral illness may have contributed to a worsening of their condition and the formal declaration of this diagnosis.

Another important phenotype identified was type 2 diabetes. Several studies have pointed out possible pathophysiological relationships between COVID-19 and diabetes.[41, 42] And the increased incidence of a number of metabolic diseases have been found with those after a COVID-19 diagnosis.[40] Our study indicates that the metabolic disorder may be so significant as to lead to a formal diagnosis of diabetes mellitus.

The disease of the nail phenotype includes a variety of diagnoses including leukonychia, onycholysis, onychomadesis, mee’s lines, muehrcke lines, and Beau’s lines all of which are markers of overall well-being and have been associated with infections, renal, or hepatic dysfunction previously. Beau’s lines have specifically been associated with COVID-19 infections.[43, 44] Our results suggest this association is widespread and likely a result of systemic infection including renal injury.

Proteinuria was also identified as having an association with COVID-19 among male patients less than 65. COVID-19 has previously been associated with acute kidney injury.[45] And proteinuria is a known surrogate for kidney disease.[46] The identification of proteinuria as an association with COVID-19 in the young patient cohort suggests the insult of COVID-19 to the kidneys persists months after the infection has resolved.

The MLHO framework appears to be more powerful than univariate PheWAS. A small number of phenotypes that had a relatively high unadjusted statistical significance (a p-value between 0.01 and 0.001) would have been dropped in a linear univariate PheWAS after p-value correction for multiple hypotheses. Two examples of such phenotypes are palpitations and non-specific chest pain, both of which have previously been described as common symptoms of PASC.[7, 35, 36]

MLHO’s implementation in this study is similar to the standard univariate PheWAS[28, 29] as both offer computational solutions for high throughput association mining from clinical data. However, a challenge in standard PheWAS is to find a sensible balance between adequately applying a correction to P-values in order to reduce false discovery due to multiple testing and minimizing false negatives.[47] Our approach expands the univariate p-value dependent criteria for identifying phenome-wide associations to a more comprehensive and multivariate entropy-based process. MLHO iteratively applies joint mutual information, performs sparsity screening, and uses gradient boosting to characterize the post-acute sequelae of COVID-19. The iterative process in MLHO provides means to an interpretable probabilistic confidence score for each phenotype associated with a past positive COVID-19 RT-PCR test.

Augmented with clinical expertise (i.e., chart reviews), MLHO’s computational algorithms avoid a flood of false-positive discoveries while offering a more robust probabilistic approach than the standard PheWAS. We were able to evaluate over 1,600 phenotypes and identify a small number of phenotypes (with confidence scores) that can indicate a past COVID-19 infection. As a result, and along with the inclusion of COVID-negative patients, this study rules out some of the phenotype associations, which were previously identified through poorly controlled observational data, such as cutaneous eruptions outside of nail changes and alopecia.

We acknowledge that this study’s findings may present limitations due to the use of only diagnosis codes, which can result in missing signs and symptoms that are in clinical notes and laboratory results. In addition, given the intensity of the pandemic and spread of misinformation, EHR data may represent confirmatory bias between providers and patients. Replicating this study in other institutions would help elucidate if the clinical phenotypes seen at MGB reflect true characteristics of PASC or local healthcare utilization patterns. Finally, we have excluded hospitalized COVID-19 patients. On the one hand, it would be difficult to match hospitalized Coronavirus patients during the COVID era with non-COVID hospitalized patients. On the other hand, the post-COVID syndrome can still be observed in patients who were never hospitalized.[12, 48–52] Regardless, future PASC studies should include hospitalized patients.

## Conclusion

The COVID-19 pandemic in the United States raged nearly uncontrolled in 2020. While the exact number of people afflicted by the post-acute sequelae of SARS-CoV-2 infection is unknown, it represents a significant public health burden because of the large magnitude of the COVID-19 spread globally. We identified 33 phenotypes that were indicative of long Covid among non-hospitalized COVID-19 patients. Our understanding of COVID-19 and its chronic sequelae is evolving and new risks are unknown. We do not know who might develop the post-COVID syndrome, how long symptoms last, and whether COVID-19 prompts the presentation of chronic diseases. Accurate identification of phenotypes will be important to guide future research and the healthcare system to focus its efforts and resources on adequately controlled age- and gender-specific sequelae of a COVID-19 infection. The ever-increasing adoption and magnitude of clinical data stored in EHR repositories over the past decade provide exceptional opportunities for instrumenting healthcare systems to study evolving pandemic byproducts. EHR data offer a unique opportunity to understand the post-acute effects that can follow SARS-CoV-2 infection.

## Data Availability

Data contains PHI and therefore is not publicly available.

## Declarations

### Ethics approval and consent to participate

The use of clinical data in this study was approved by the MGB Institutional Review Board with a waiver of informed consent

### Availability of data and materials

The anonymized patient-level data used for this project cannot be shared for reasons of information governance. Data may be available to affiliated researchers given the MGB IRB approval.

## Acknowledgments

We thank many colleagues in the Mass General Brigham Research Information Science & Computing team, for curating MGB COVID-19 mart and providing information science and computing support.

